## Supplementary Information for "Ultrafast topological data analysis reveals pandemic-scale dynamics of convergent evolution"

Michael Bleher[1*†], Lukas Hahn[1†], Maximilian Neumann[1,2*†], Zachary Ardern[7], Juan Ángel Patiño-Galindo [4], Mathieu Carrière[5], Ulrich Bauer[6], Raúl Rabadán[3*], Andreas Ott[1*]

[1]Institute for Mathematics, Heidelberg University, Heidelberg, Germany

[2]Institute for Biological Interfaces 5, Microbial Genetics & Biotechnology, Karlsruhe Institute of Technology, Karlsruhe, Germany

[3]Program for Mathematical Genomics, Department of Systems Biology, Columbia University, New York, NY, USA

[4]Department of Microbiology, Icahn School of Medicine at Mount Sinai, New York, NY, USA

[5]DataShape, Centre Inria d'Université Côte d'Azur, Biot, France

[6]TUM Department of Mathematics and Munich Data Science Institute, Munich, Germany

[7]Wellcome Trust Sanger Institute, Hinxton, United Kingdom

[†]These authors contributed equally to this work.

[*]Corresponding authors:
 (M.B.)
 (M.N.)
 (R.R.)
 (A.O.)

### 1 Acquisition and preparation of sequence alignments

#### 1.1 Coronavirus

We used SARS-CoV-2 genome data shared via the GISAID EpiCoV Database, the global data science initiative [1], and accessible at https://doi.org/10.55876/gis8.240314pc. We downloaded the alignment msa_0222.fasta on 24 February 2024. This alignment comprises 14,956,592 SARS-CoV-2 nucleotide sequences with collection dates between December 2019 and February 2024 that were aligned to the reference sequence hCoV-19/Wuhan/WIV04/2019 (EPI_ISL_402124) with MAFFT v7.497 [2]. We generated the following sub-alignments:

##### 1.1.1 SARS-CoV-2 whole genome alignment

We applied the following operations to the alignment msa_0222.fasta:

(i) Sequences were truncated to the whole genome (nucleotide positions 266 to 29,674 in the reference sequence hCoV-19/Wuhan/WIV04/2019).
(ii) All sequences containing any letters other than A, C, T, G or - were removed.
(iii) All sequences that did not occur in at least three identical copies in the resulting alignment were removed.
(iv) Among each set of identical sequences, all but the earliest sequence (based on collection date) were removed.
(v) Sequences in the alignment were arranged in time-reversed order (by collection date).

This resulted in an alignment with 283,687 distinct whole genomes of length 51,395nt covering the period from December 2019 until February 2024.

##### 1.1.2 SARS-CoV-2 spike gene alignment

We applied the following operations to the alignment msa_0222.fasta:

(i) Sequences were truncated to the spike gene (nucleotide positions 21,563 to 25,384 in the reference sequence hCoV-19/Wuhan/WIV04/2019).
(ii) All sequences containing any letters other than A, C, T, G or - were removed.
(iii) All sequences that did not occur in at least three identical copies in the resulting alignment were removed.
(iv) Among each set of identical sequences, all but the earliest sequence (based on collection date) were removed.
(v) Sequences in the alignment were arranged in time-reversed order (by collection date).

This resulted in an alignment with 158,190 distinct spike genes of length 8,237nt covering the period from December 2019 until February 2024.

##### 1.1.3 SARS-CoV-2 Omicron spike gene alignment

We applied the following operations to the spike gene alignment (see Section 1.1.2):

(i) All sequences that share at least 50% of the defining spike amino acid changes of the Omicron lineages BA.1, BA.2 and XBB.1.5 [3] were selected. Here the defining spike amino acid changes of the Omicron lineages were taken to be G142D, G339D, S371L, S371F, S373P, S375F, K417N, N440K, S477N, T478K, E484A, Q493R, Q498R, N501Y, Y505H, D614G, H655Y, N679K, P681H, N764K, D796Y, Q954H, N969K.
(ii) All sequences with collection date before November 2021 (date of discovery of the Omicron variant) were removed.

(iii) An Omicron spike gene reference sequence was created by introducing the following single nucleotide substitutions (corresponding to the amino acid changes listed in (i)) to the reference sequence hCoV-19/Wuhan/WIV04/2019 (EPI_ISL_402124): G21987A (G142D), G22578A (G339D), T22673C (S371P), C22674T (S371F), T22679C (S373P), C22686T (S375F), G22813T (K417N), T22882G (N440K), G22992A (S477N), C22995A (T478K), A23013C (E484A), A23040G (Q493R), A23055G (Q498R), A23063T (N501Y), T23075C (Y505H), A23403G (D614G), C23525T (H655Y), T23599G (N679K), C23604A (P681H), C23854A (N764K), G23948T (D796Y), A24424T (Q954H), T24469A (N969K). This reference sequence was added to the alignment.

This resulted in an alignment with 72,309 distinct spike genes of length 8,237nt covering the period from November 2021 until February 2024.

##### 1.1.4 SARS-CoV-2 United Kingdom spike gene alignment

We applied the following operations to the alignment msa_0222.fasta:

(i) Sequences were truncated to the spike gene (nucleotide positions 21,563 to 25,384 in the reference sequence hCoV-19/Wuhan/WIV04/ 2019).

(ii) All sequences containing any letters other than A, C, T, G or - were removed.

(iii) All sequences that were not from the United Kingdom (England, Wales, Scotland, Northern Ireland) were removed.

(iv) All sequences with collection date after August 2022 were removed.

(v) All sequences that did not occur in at least three identical copies in the alignment were removed.

(vi) Among each set of identical sequences, all but the earliest sequence (based on collection date) were removed.

(vii) Sequences in the alignment were arranged in time-reversed order (by collection date).

(viii) The reference sequence hCoV-19/Wuhan/WIV04/2019 (EPI_ISL_402124) was added to the alignment.

This resulted in an alignment with 25,713 distinct spike genes of length 8,237nt covering the period from December 2019 until August 2022.

##### 1.1.5 SARS-CoV-2 alignments for benchmarking

We created eight SARS-CoV-2 sequence alignments by sorting the SARS-CoV-2 whole genome alignment (see Section 1.1.1) according to collection date in ascending order, and then selecting the first $n$ sequences for $n = 1{\times}10^3, 2{\times}10^3, 5{\times}10^3, 1{\times}10^4, 2{\times}10^4, 5{\times}10^4, 1{\times}10^5, 2{\times}10^5$. In each of these alignments, the first sequence is the reference sequence hCoV-19/Wuhan/WIV04/2019 (EPI_ISL_402124).

##### 1.1.6 SARS-CoV-2 whole genome alignment for sequencing error analysis

We applied the following operations to the alignment msa_0222.fasta:

(i) Sequences were truncated to the whole genome (sites 266 to 29,674 in the reference sequence hCoV-19/Wuhan/WIV04/2019).

(ii) All sequences containing any letters other than A, C, T, G or - were removed.

(iii) The first 200,000 sequences in the resulting alignment were selected.

##### 1.2 Avian influenza virus

We used avian influenza A subtype H5N1 (strain 2.3.4.4b) genome datasets for the HA and PB2 genes, provided by the GISAID EpiFlu Database [1] and accessible at `https://doi.org/10.55876/gis8.251116eh` (HA gene) and `https://doi.org/10.55876/gis8.251116bc` (PB2 gene). For each gene we downloaded all available nucleotide sequences (complete, with complete collection date not before 30 December 2021) as of 16 November 2025. Subsequently we aligned sequences separately in each of the resulting datasets to the reference sequence A/American_Wigeon/South_Carolina/22-000345-001/2021 (EPI_ISL_18133029) using MAFFT v7.526 [2] with arguments `--thread -1 --anysymbol --auto`. We generated the following two alignments:

###### 1.2.1 Influenza H5N1 HA gene alignment

We downloaded and aligned 24,141 sequences, and applied the following operations:

(i) All sequences containing any letters other than A, C, T, G or - were removed.
(ii) Data augmentation (ancestral sequence reconstruction) was applied (see Section 1.4).
(iii) Duplicate sequences were removed.
(iv) Among each set of identical sequences, all but the earliest sequence (based on collection date) were removed.

This resulted in an alignment with 17,223 distinct HA genes of length 2,571nt covering the period from December 2021 until November 2025.

###### 1.2.2 Influenza H5N1 PB2 gene alignment

We downloaded and aligned 23,237 sequences, and applied the following operations:

(i) All sequences containing any letters other than A, C, T, G or - were removed.
(ii) Data augmentation (ancestral sequence reconstruction) was applied (see Section 1.4).
(iii) Duplicate sequences were removed.
(iv) Among each set of identical sequences, all but the earliest sequence (based on collection date) were removed.

This resulted in an alignment with 18,139 distinct PB2 genes of length 2,357nt covering the period from December 2021 until November 2025.

##### 1.3 Human immunodeficiency virus Env gene alignment

We used HIV-1 (strain HXB2) genome data provided by the European Nucleotide Archive (ENA) [4] (for accession numbers see Supplementary Table 4). We downloaded 30,964 envelope (Env) gene nucleotide sequences with genome coverage of at least 78% on 1 March 2025 and aligned them to the HIV1/HTLV-III/LAV reference genome (ENA accession number K03455.1) using MAFFT v7.526 [2] with arguments `--thread -1 --anysymbol --auto`. We applied the following operations to the resulting alignment:

(i) All sequences containing any letters other than A, C, T, G or - were removed.
(ii) Data augmentation (ancestral sequence reconstruction) was applied (see Section 1.4).
(iii) Duplicate sequences were removed.

This resulted in an alignment with 30,088 distinct Env genes of length 36,265nt.

##### 1.4 Data augmentation

To compensate for the low sampling rate in the H5N1 and HIV-1 alignments, we applied data augmentation via ancestral sequence reconstruction. We added synthetic sequences such that all edges of Hamming length 2 or 3 in the complete geometric graph of the original alignment were replaced by chains of unit-length edges (see Supplementary Figure 2a-b). Given an edge connecting two sequences, we first determined the nucleotide positions at which the two sequences differed and then generated synthetic sequences by sequentially replacing the nucleotides in the second sequence with those from the first sequence at these positions.

In alignments including collection date, synthetic sequences constructed from a pair of sequences in the alignment were all assigned the date of the most recent of these two sequences. We incorporated collection dates in the data augmentation for the H5N1 alignments, but we did not do so for the HIV-1 alignment.

This deterministic procedure of ancestral sequence reconstruction may introduce systematic bias, as the outcome depends on the specific order in which nucleotide substitutions are performed. To quantify this bias, we randomized the procedure by randomly permuting the order of nucleotide substitutions. We then compared the tRI profiles of alignments generated by the randomized procedure with those obtained from the deterministic procedure by computing Pearson correlation coefficients (see Supplementary Figure 2c).

#### 2 Statistical analyses

##### 2.1 Topological signals in neutral evolution

Topological cycles can arise randomly in neutral evolution, resulting in some background noise in tRI. To estimate this topological background noise, we analyzed topological signals (topological cycles, SNV cycles and tRI) in simulated neutral evolutionary scenarios which we generated under the following assumptions: uniform probability distribution for substitutions across the genome, no variations in fitness, and zero recombination rate.

Simulations were based on a Wright-Fisher forward model of viral evolution and performed with SANTA-SIM v1.0 [5] with fixed parameters: genome length ($30,000$nt), number of generations ($N = 10,000$), number of sequences sampled from the population per time step ($n = 15$), recombination rate ($\rho = 0$), transition bias (1.0) and variable parameters: mutation rate per site per generation $\mu$, population size $p$, carrying population $c$, population growth rate per generation $g$. Each simulation was initiated with a population of $p$ polyadenine sequences AA. . . A of length $30,000$nt. We considered five scenarios: in scenarios I-III we varied the mutation rate $\mu$ under the assumption of fixed population size $p$, while in scenarios IV and V we investigated the effects of logistic growth $g$ of the viral population with carrying population $c$ (see Supplementary Figure 5a-b). We removed duplicate sequences from the resulting simulated alignments to obtain alignments with distinct sequences (which may then have varying number of sequences).

The range of mutation rates in scenarios I-III was chosen such that the diversity in the simulated phylogenies is in close correspondence to the observed diversity in the SARS-CoV-2 200k whole genomes alignment used for benchmarking (see Section 1.1.5). While a mutation rate of $\mu = 0.75 \times 10^{-7}$ substitutions per generation per site underestimates the maximal distances to the root, the highest value of $\mu = 1.25 \times 10^{-7}$ produces slightly larger maximal values. Scenario II with an intermediate value of $\mu = 1.00 \times 10^{-7}$ reproduced the observed maximal distances accurately and therefore provides a good approximation of the SARS-CoV-2 whole genome alignment.

##### 2.1.1 Topological cycles

In order to keep overall computational expenses at a reasonable level, we resorted to extrapolations from smaller simulated datasets to the size of the SARS-CoV-2 200k whole genomes alignment. For all scenarios we produced 100 simulations for each of the following values of the population size $p$ (resp. carrying population $c$): 100, 500, 1000, 2500, 5000, 7500, $10^4$, $10^5$. Additionally, we included five simulations for $p = 10^6$ to achieve a better support of the extrapolation fit at large values. We computed the full persistent homology in dimension one (all persistence barcodes and topological cycles) of the simulated alignments using Hammingdist v1.4.0 [6] and Ripser [7, 8] (see Supplementary Figure 5c and Methods). For each value of $p$ we randomly chose 60% of the simulations as training data, used to determine the parameters of different models in a non-linear least squares fit, while the remaining 40% were reserved for later validation and comparison of the models.

For each scenario we considered a quadratic, cubic, powerlaw and exponential model for the observed points $(x_i, y_i)$, and linear and powerlaw fits for the squared residuals $(y_i - y_{\text{fit}}(x_i))^2$ in the training data (see Figure 2). In each model, we then used the resulting fits mean$(x)$ and var$(x)$ as estimators for the mean and variance of an underlying Panjer $(a, b, 0)$-class distribution, a class of distributions that encompasses the binomial, Poisson, and negative binomial distribution [9, 10]. The quantiles of the observed number of cycles in the training data fit the quantiles of the Panjer distribution with corresponding mean and variance remarkably well (see Figure 1). We then determined the likelihood $L = \prod_i P_{\text{Panjer}}(y = y_i | \text{mean}(x_i), \text{var}(x_i))$ to observe the validation data $\{(x_i, y_i)\}$. For each model, the corresponding log-likelihoods are listed alongside the corresponding fits in Figure 2. According to the log-likelihoods, the variance of the Panjer distribution is generally best described by a powerlaw behaviour. An exception is scenario II, for which the small sample of 5 simulations at $p = 10^6$ has an uncharacteristically small variance that skews the fits and corresponding likelihoods. Among the models that assume a powerlaw dependence of the variance, again with exception of scenario II, the cubic-powerlaw model yields maximum likelihoods.

Based on these results, we computed the 95% prediction intervals for the expected numbers of random topological cycles using a cubic extrapolation for the mean and a powerlaw extrapolation for the variance of a Panjer distribution. The validation data of scenarios I, IV and V, which were all based on the same mutation rate, are well described by the prediction intervals of scenario I. While the prediction intervals of scenario V differ significantly from the other two scenarios at high numbers of distinct sequences, this difference arises only because simulations in scenario V generally produce fewer distinct sequences than scenario I and IV, such that a steeper extrapolation is not sufficiently penalized. Hence, the prediction intervals of scenario V illustrate the error margins of the extrapolations, but are not likely to faithfully represent the expected number of topological cycles. We also observed that higher mutation rates in scenarios II and III lead to smaller numbers of SNV cycles in the dataset. Since the diversity of the SARS-CoV-2 alignment is arguably better approximated by scenario II than by scenarios I, III, IV or V, it is reasonable to rely on the prediction intervals of scenario II.

##### 2.1.2 tRI signals and SNV cycles

For each of the scenarios I-V above, we ran 10 independent simulations with the following parameters: population size $p = 10^6$ for scenarios I, II, and III; $p = 100$ and carrying population $c = 10^6$ for scenarios IV and V. The resulting alignments comprised 150k sequences. We removed duplicate sequences and performed a topological recurrence analysis with EVOtRec for each of the resulting alignments, and also for the SARS-CoV-2 50k and 100k whole genomes alignments used for benchmarking (see Section 1.1.5). We subsequently computed summary statistics of topological signals for each alignment, including the total number of SNV cycles, the total number of SNVs with (positive) tRI signal, the total sum of tRI signals and the maximum tRI signal (see Figure 2a and Supplementary Figure 5d). In these analyses, the tRI was obtained from the raw tRI by subtracting one tRI unit (see Methods), but no normalization using four-fold degenerate sites was applied as alignments contain simulated sequences.

#### 2.2 Robustness of tRI to stochastic sequencing errors

To analyze the effect of stochastic sequencing errors on tRI, we added simulated stochastic sequencing errors to nucelotide sequence alignments and computed tRI signals with EVOtRec for the original alignments and for the alignments with added sequencing errors. We performed two such analyses, based on two reference alignments: a synthetic alignment generated with SANTA-SIM v1.0 [5] for the neutral evolutionary scenario II described in Section 2.1, and a specially prepared real SARS-CoV-2 200k whole genomes alignment (see Section 1.1.6). We added simulated sequencing errors to each of these alignments with the following error rates per nucleotide site: $10^{-3}$, $10^{-4}$, $10^{-5}$ and $10^{-6}$ for the scenario II alignment, and $10^{-3}$, $10^{-4}$, $9 \times 10^{-5}$, $\ldots$, $2 \times 10^{-5}$, $10^{-5}$, $10^{-6}$, $10^{-7}$, $10^{-8}$ for the SARS-CoV-2 alignment. Here at every position on the sequence with nucleotide A, C, T or G, this nucleotide was replaced by a randomly selected different nucleotide with probability equal to the error rate. We ran 10 independent simulations for each of the given error rates. We subsequently removed duplicate sequences from the resulting alignments to obtain alignments with distinct sequences (these alignments may therefore contain varying numbers of sequences) and performed a topological recurrence analysis with EVOtRec.

To quantify the absolute effect of stochastic sequencing errors on tRI, we computed summary statistics of tRI signals across both analyses. Specifically, we evaluated the number of SNV cycles, the number of SNVs with tRI signal, the total sum of tRI signals and the maximum tRI signal (see Figure 2b and Supplementary Figure 6). In these analyses, the tRI was obtained from the raw tRI by subtracting one tRI unit (see Methods), but no normalization using four-fold degenerate sites was applied as alignments contain simulated sequences.

To quantify the relative effect of stochastic sequencing errors on tRI signals in the SARS-CoV-2 alignment, we computed the Pearson correlation between tRI profiles across the whole genome (i.e. list of observed tRI signals for all SNVs in the alignment) for alignments with added sequencing errors vs. the original alignment with no added sequencing errors (reference alignment) (see Supplementary Figure 6b2). Since the alignments associated with sequencing error rate $10^{-3}$ all exhibited no tRI signals in our analysis, the corresponding Pearson correlation is not well-defined in this case.

To mitigate the effect of stochastic sequencing errors on tRI we applied the following filtering rules in the preprocessing of sequence alignments (see Section 1):

(i) Sequences containing any letters other than A, C, T, G or - were removed.

(ii) Only complete sequences of high-quality were retained.

(iii) Sequences that do not occur in at least three identical copies in the alignment were removed.

We relaxed condition (iii) for the H5N1 alignments and conditions (ii) and (iii) for the HIV alignment, because the sampling rate in the original alignments was too low for a meaningful topological recurrence analysis (see Section 1).

#### 2.3 Monte Carlo permutation test

We compared frequency distributions of raw tRI signals in the SARS-CoV-2 whole genome and spike gene alignments with simulated frequency distributions of raw tRI signals under the assumption that raw tRI signals are distributed uniformly across the genome (see Supplementary Figure 7). First, we computed raw tRI frequency distributions for the SARS-CoV-2 whole genome and spike gene alignments. Next we simulated raw tRI frequency distributions. For this, we computed the total sum of raw tRI signals for the whole genome alignment (spike gene alignment). These signals were then randomly distributed across nucleotides on the whole genome (spike gene) with a uniform probability distribution. We ran 100 independent iterations of this simulation and took means per nucleotide position to obtain the final simulated raw tRI frequency distributions. Finally, we used a Kolmogorov–Smirnov test to compare these simulated raw tRI frequency distributions with the corresponding real raw tRI frequency distributions.

#### 2.4 Significance of tRI signals

To evaluate whether the tRI signal of a single nucleotide variant (SNV) is statistically significant (i.e. higher than expected by chance), we computed a conservative significance level for tRI signals resulting from a given sequence alignment. The sequence alignment determines a genomic region which it covers. We first implemented a permutation test that compares the observed raw tRI signals to a baseline in which all possible SNVs in the genomic region are assumed to contribute to tRI signals with equal probability. This null hypothesis corresponds to a multinomial distribution of tRI values across all possible SNVs in the genomic region, so we can compute its exact tail analytically without resampling.

Formally, we assigned a numbering $1, 2, \ldots, 3L-1, 3L$ to all possible SNVs in the genomic region, where $L$ is the number of nucleotide sites in the genomic region. The number of possible SNVs is $3L$, since every site admits three possible nucleotide substitutions. Each number $s$ in this numbering corresponds to a unique SNV and vice versa. Let the nonnegative integer $k_s \in \mathbb{Z}_{\geq 0}$ denote the raw tRI of a given SNV $s$ and let $N = \sum_{s=1}^{3L} k_s$ be the total number of observed raw tRIs in the sequence alignment. Under the null hypothesis, the joint distribution of raw tRI signals is multinomial with equal cell probabilities, hence for any fixed $s$ we have $k_s \sim \mathrm{Bin}\big(N, \frac{1}{3L}\big)$. Given an observed raw tRI $k = k_s$, the one-sided $p$-value is the binomial upper tail

$$p(k \mid N, 3L) = \Pr\left[\mathrm{Bin}\left(N, \frac{1}{3L}\right) \geq k\right] = \sum_{j=k}^{N} \binom{N}{j} \left(\frac{1}{3L}\right)^{j} \left(1 - \frac{1}{3L}\right)^{N-j}.$$

The tRI significance level is the integer $K_\alpha(N, 3L)$ for which $p(K_\alpha \mid N, 3L) < \alpha$, i.e.

$$K_\alpha(N, 3L) = F^{-1}_{N,1/(3L)}(1-\alpha) = \min\{\, k \in \mathbb{Z}_{\geq 0} : F_{N,1/(3L)}(k) \geq 1 - \alpha \,\},$$

where $F_{N,1/(3L)}$ is the cumulative distribution function of $\mathrm{Bin}\big(N, \frac{1}{3L}\big)$. We used the usual $p$-value bound $\alpha = 0.05$ throughout this study. We remark that under the null hypothesis, we have $\mathbb{E}[k_s] = N/(3L)$ and $\mathrm{Var}(k_s) = \frac{N}{3L}\big(1 - \frac{1}{3L}\big)$. In fact, a Poisson approximation with rate $\lambda = N/(3L)$ can be used when $L$ is large (and $\lambda$ moderate), although all thresholds and $p$-values reported in this study use the exact binomial tail.

From this significance level for the raw tRI we computed a normalized significance level for the actual tRI. For this, we rescaled the raw tRI significance level for each nucleotide mutation type, by dividing the the raw tRI significance level by the corresponding raw tRI mean per nucleotide mutation type across four-fold degenerate sites (see Methods). If this average happened to be zero, the rescaling was not well-defined and no rescaling was applied. In the case of time series tRI data, the tRI significance level was computed at every time step. We note that this may lead to spurious significance levels at the onset of positive rescaling factors.

Our definition of the tRI significance level is based on the null hypothesis that all possible SNVs in the genomic region contribute to tRI signals with equal probability. The significance level is conservative because it does not distinguish between signal and noise.

##### 2.5 Effect of the ambiguity of cycle representatives and bases on tRI

The computation of (raw) tRI signals via exhaustive reduction with Ripser (see Methods) provides an efficient way to extract persistence features (over time). This amounts to choosing a set of cycle representatives that live in the cycle space over $\mathbb{F}_2$ of the underlying unit distance graph in the Vietoris-Rips complex. Let $C = (c_1, \ldots, c_r)$ be a collection of such (exhaustive) cycle representatives. Then we can extend $C$ to an $\mathbb{F}_2$-basis of the full cycle space by adding a collection of triangles $(c_{r+1}, \ldots, c_k)$. Now given a mutation XposY (“X”, “Y” denote nucleotides and “pos” the position on the genome) and the collection of all unit-length edges $(e_1, \ldots, e_m)$ that contain XposY, we can form an $m \times (r+k)$ matrix $M = M(\text{XposY})$ over $\mathbb{F}_2$ where $M_{i,j} = 1$ if $c_j$ contains $e_j$ and $M_{i,j} = 0$ if not.

To study the effect of basis changes in persistent homology on the tRI, we need to address the following two points. First, we have to understand how column operations among the representatives $(c_1, \ldots, c_r)$ affect the tRI. Suppose that we add a column $c_i$ to column $c_j$ for $i, j \in \{1, \ldots, r\}$ with $i \neq j$. If both columns $M_{c_i}, M_{c_j} = 0$, the tRI obviously remains unchanged. Here we denote by $M_c$ the column in $M$ corresponding to the cycle $c$. The following scenarios remain:

1. $M_{c_i}, M_{c_j} \neq 0$ and $M_{c_i} + M_{c_j} \neq 0$,
2. $M_{c_i}, M_{c_j} \neq 0$ and $M_{c_i} + M_{c_j} = 0$,
3. $M_{c_i} \neq 0, M_{c_j} = 0$ and $M_{c_i} + M_{c_j} \neq 0$.

Once we counted an edge $e_j$ in a cycle $c_i$, the row in $M$ corresponding to $e_j$ will be deleted and contributes to the raw count of the mutation XposY. This raw count is obviously independent of the ordering of the cycle representatives and unchanged in the given scenarios 1-3. For the computation of the (raw) tRI, once we counted the contribution of an edge $e_j$ in a cycle $c_i$ to the raw count of the mutation XposY, we additionally delete all other rows where $M_{i,k} = 1$ to avoid double counting. We observed that on average, this changes the magnitude of (raw) tRI signals across all alignments (see Figure 3). Moreover, we found a strong correlation between raw counts and raw tRI in SNV cycles.

Second, an additional level of ambiguity arises from the fact that adding triangles does not change the homology class of a given cycle representative. To estimate the potential information loss, we counted edges that contain XposY and participate in triangles but which are not contained in $(c_1, \ldots, c_r)$ (see Figure 4). We found that the potential effect on tRI signals across our datasets is overall small.

#### 3 Validation on simulated data

##### 3.1 The effect of fitness and sampling density on tRI in simulated data

We investigated the effects on raw tRI signals of two key parameters in genome evolution: fitness and sampling density of the underlying phylogeny (see Figure 2c-d and Supplementary

Figure 3). For this, we used simulations of viral evolution generated with SANTA-SIM v1.0 [5] (see Section 2.1).

Each simulation was initialized with a population of 100k polyadenine sequences AA. . . A of length 2000nt. The base setting for these simulations was chosen as follows: purely clonal replication, excluding recombinations; a uniform neutral fitness parameter of 1.0 for all nucleotide mutations; a mutation rate of $1.00 \times 10^{-5}$ variations per nucleotide per generation; a transition bias of 1.0. Every simulation was run for a total of 2000 generations, where in each generation we collected 20 samples. The resulting alignments comprised 40k sequences.

We systematically varied one of the two parameters fitness or sampling density, while keeping the other parameter fixed. The scenarios were as follows:

- Fitness: The fitness parameter assigned to C alleles at the first 100 nucleotide positions (variants with varied fitness) was uniformly set to one of the values 0.98, 1.00, 1.02, 1.04, 1.06, 1.08 and 1.10, making the mutations A1C, A2C, . . . , A100C deleterious, neutral or beneficial, respectively. Fitness parameters in this range are typical for single nucleotide mutations in RNA viruses [11]. The fitness parameter for all other mutations was kept at 1.00 (neutral variants). For each of these scenarios we ran 20 independent simulations.
- Sampling density: We set the fitness parameter assigned to C alleles at the first 100 nucleotide positions uniformly to the level 1.10 (beneficial variants) and kept it at 1.00 for all other mutations (neutral variants). We ran 20 independent simulations and from each simulated alignment, we drew random subsamples comprising 10%, 20%, 30%, 40%, 50%, 60%, 70%, 80%, 90% and 100% of sequences.

We removed duplicate sequences from the resulting alignments and performed a topological recurrence analysis with EVOtRec. For each scenario, we computed the mean tRI per SNV separately by averaging across variants with varied fitness (the 100 SNVs A1C, A2C, . . . , A100C) and by averaging across neutral variants. In these analyses, the tRI was obtained from the raw tRI by subtracting one tRI unit (see Methods), but no tRI normalization using four-fold degenerate sites was applied as alignments contain simulated sequences.

##### 3.2 Comparison with phylogenetic homoplasy counts in simulated data

We compared EVOtRec's raw tRI signals with a tree-based count of homoplasies (see Supplementary Figure 4), using simulated sequence alignments generated with SANTA-SIM v1.0 [5] (see Section 2.1).

Each simulation was initialized with a fixed population of 100k polyadenine sequences AA. . . A of length 1000nt. The parameters for the simulations were chosen as follows: purely clonal replication, excluding recombinations; a fitness parameter of 1.3 assigned to C alleles at the first 200 nucleotide positions (beneficial variants), while all other mutations were assigned a fitness parameter of 1.00 (neutral variants); a mutation rate of $1.00 \times 10^{-5}$ variations per nucleotide per generation; a transition bias of 1.0. Every simulation was run for a total of 2000 generations, where in each generation we collected 10 sequences. The resulting alignments comprised 20k sequences. We ran 100 independent simulations.

For each simulated alignment, we removed duplicate sequences and computed raw tRI signals with EVOtRec. For comparison with phylogenetic homoplasy counts, we furthermore constructed a phylogenetic tree for each simulated alignment using IQ-TREE v3.0.1 [12, 31] with arguments `-m HKY --fast`, and then performed a homoplasy count analysis using TreeTime v0.11.4 [13] with arguments `homoplasy -n 2000`. Parameters in simulations with SANTA-SIM were chosen in such a way that the resulting alignments were of a moderate size which IQ-TREE could handle in reasonable time.

Next, we computed mean raw tRI signals and mean TreeTime homoplasy counts per SNV, where means per SNV were calculated by averaging over all 100 simulations. We also determined the mean Spearman correlation between raw tRI profiles and TreeTime homoplasy counts, separately across neutral variants and across beneficial variants, where means $\pm$ 95% confidence interval (CI) were calculated by averaging over all 100 simulations.

#### 4 Validation on experimental data

We validated tRI signals of amino acid changes in the SARS-CoV-2 (Omicron) spike gene alignments and in the H5N1 HA and PB2 gene alignments (see Section 1) on published deep mutational scanning (DMS) data. We extracted tRI/prevalence data from Supplementary Tables 2 and 3. We subsequently aggregated tRI/prevalence signals of non-synonymous SNVs into tRI/prevalence signals of SAAVs by summing up the tRI/prevalence signals of all SNVs that give rise to the same SAAV.

##### 4.1 SARS-CoV-2 spike gene

We computed the time-dependent Pearson correlation between SARS-CoV-2 spike gene tRI and DMS effect [14] (column "delta_bind" under the condition "target = Wuhan-Hu-1" in [15]) for amino acid changes on the wild-type receptor-binding domain (RBD), for every month between May 2020 and December 2023. We assigned tRI = 0 to SAAVs for which no tRI signal was available. The monthly Pearson correlation was then computed on the set of RBD amino acid changes for which (i) the monthly prevalence was positive and (ii) both tRI data and DMS data were available.

##### 4.2 SARS-CoV-2 Omicron spike gene

We compared tRI signals in the SARS-CoV-2 Omicron spike gene alignment with DMS effects for the BA.1 sub-lineage [16] (column "effect" in [17]), the BA.2 sub-lineage [18] (column "spike mediated entry" in [19]) and the XBB.1.5 sub-lineage [18] (columns "ACE2 binding", "spike mediated entry" and "human sera escape" in [19]). We assigned tRI = 0 to SAAVs for which no tRI signal was available. For each DMS effect, we computed a $2 \times 2$ contingency table for binarized tRI (tRI $> 0$ vs. tRI $= 0$) and binarized DMS effect (DMS effect $> 0$ vs. DMS effect $\leq 0$). Based on this table, we performed a Fisher's exact test and computed precision, recall, $F_1$ score and Phi coefficient. Contingency tables were computed on the set of all SAAVs for which both tRI data and all DMS data per sub-lineage were available. In addition, for the XBB.1.5 sub-lineage we computed a logistic regression model predicting positive tRI as a function of the three XBB.1.5 DMS effects.

We repeated all analyses with tRI replaced by published phylogeny-based fitness effect estimates from [20] (BA.1: column "delta_fitness" under the conditions "gene = S" and "clade = 21K" in [21]; BA.2: column "delta_fitness" under the conditions "gene = S" and "clade = 21L" in [21]; XBB.1.5: "delta_fitness" under the conditions "gene = S" and "clade = 23A" in [21]) and [22] (BA.1: column "delta_fitness" under the condition "gene = S" in [23]; BA.2: column "delta_fitness" under the condition "gene = S" in [24]; XBB.1.5: column "delta_fitness" under the condition "gene = S" in [25]). For the XBB.1.5 sub-lineage we computed an ordinary least squares regression model predicting positive fitness effect as a function of the three XBB.1.5 DMS effects.

##### 4.3 Influenza H5N1 HA and PB2 genes

We compared tRI signals in the H5N1 HA and PB2 gene alignments with DMS effects for the HA gene [26] (column "entry in 293T cells" in [27]) and PB2 gene [28] (column "log2effectA549" in [29]). We assigned tRI $= 0$ to SAAVs for which no tRI signal was available. For each alignment, we computed a $2 \times 2$ contingency table for binarized tRI (tRI $> 0$ vs. tRI $= 0$) and binarized DMS effect (DMS effect $> 0$ vs. DMS effect $\leq 0$). Based on this table, we performed a Fisher's exact test and computed precision, recall, $F_1$ score and Phi coefficient. Contingency tables were computed on the set of all SAAVs for which (i) the prevalence was positive and (ii) both tRI data and all DMS data were available.

#### 5 Performance analysis and benchmarking

We analyzed the runtime and peak memory usage (RAM) of EVOtRec and four phylogenetic tree reconstruction tools on eight SARS-CoV-2 whole genome alignments, denoted by `alignment_1.fasta`, ..., `alignment_8.fasta`, containing $1 \times 10^3$, $2 \times 10^3$, $5 \times 10^3$, $1 \times 10^4$, $2 \times 10^4$, $5 \times 10^4$, $1 \times 10^5$ and $2 \times 10^5$ distinct sequences (see Section 1.1.5). Each tool was run on all alignments for which the runtime did not exceed 24 hours. We measured the runtime and peak RAM usage (MaxRSS) with GNU time v1.7. Subsequently, for each alignment we computed the mean $\pm$ 95% CI from measurements obtained over 3 independent runs. All computations were performed on a server with Intel Xeon Gold 6230R processors (2.10GHz) and 52 cores.

##### 5.1 EVOtRec

We performed a de novo topological recurrence analysis with the EVOtRec pipeline for for each of the alignments `alignment_1.fasta`, ..., `alignment_8.fasta` with up to 200k sequences. We used the Python script evotrec.py v1.0, available via `https://github.com/ottamj/evotrec`. We ran evotrec.py with default arguments (no time series analysis) in PyPy v3.9. For better performance, we reordered the sequence alignments in descending order based on collection date before running the analysis (see Methods and [30]).

##### 5.2 IQ-TREE

We performed a de novo tree reconstruction with IQ-TREE v3.0.1 [12, 31] for the alignments `alignment_1.fasta`, ..., `alignment_5.fasta` with up to 20k sequences. For each alignment we ran IQ-TREE with arguments `-s alignment_n.fasta -m HKY -redo -fast`.

##### 5.3 VeryFastTree

We performed a de novo tree reconstruction with VeryFastTree v4.0.5 [32, 33] for the alignments `alignment_1.fasta`, ..., `alignment_8.fasta` with up to 200k sequences. For each alignment we ran VeryFastTree with arguments `-gtr -nt -threads 52 -threads-mode 1 -noml -fastest alignment_n.fasta`.

##### 5.4 CMAPLE

We performed a de novo tree reconstruction with CMAPLE v1.1.0 [34, 35] for each of the alignments `alignment_1.fasta`, ..., `alignment_6.fasta` with up to 50k sequences. For each alignment we ran CMAPLE with arguments `-aln alignment_n.fasta --search FAST`.

##### 5.5 UShER

We performed an iterative tree reconstruction with UShER v0.6.6 [36] for the alignments `alignment_1.fasta`, ..., `alignment_8.fasta` with up to 200k sequences. Initially, the alignment `alignment_1.fasta` was converted into a VCF file `alignment_1.vcf` using UShER's tool faToVcf. Then the phylogenetic tree `alignment_1.treefile` that had previously been constructed from the alignment `alignment_1.fasta` with IQ-TREE (see subsection 5.2), and the VCF file `alignment_1.vcf` were converted into a mutation-annotated tree protobuf file `alignment_1.pb` using UShER's preprocessing mode by running UShER with arguments `-t alignment_1.treefile -v alignment_1.vcf -o alignment_1.pb`. Subsequently, for all other alignments `alignment_2.fasta`, ..., `alignment_8.fasta` the following two-step iterative process was applied:

(i) The alignment `alignment_n.fasta` was converted into a VCF file `alignment_n.vcf` using UShER's tool faToVcf.

(ii) Sequences from the alignment `alignment_n.fasta` were placed onto the existing tree that had previously been constructed from `alignment_n-1.fasta`, by running UShER with arguments `-i alignment_n-1.pb -v alignment_n.vcf -o alignment_n.pb`.

**Figure 1. Quantile-quantile analysis of Panjer distribution versus observed number of one-dimensional cycles in simulated phylogenies.** For each value of carrying population $c$ we determined the mean and variation of the observations and used these as parameters for the Panjer distribution.

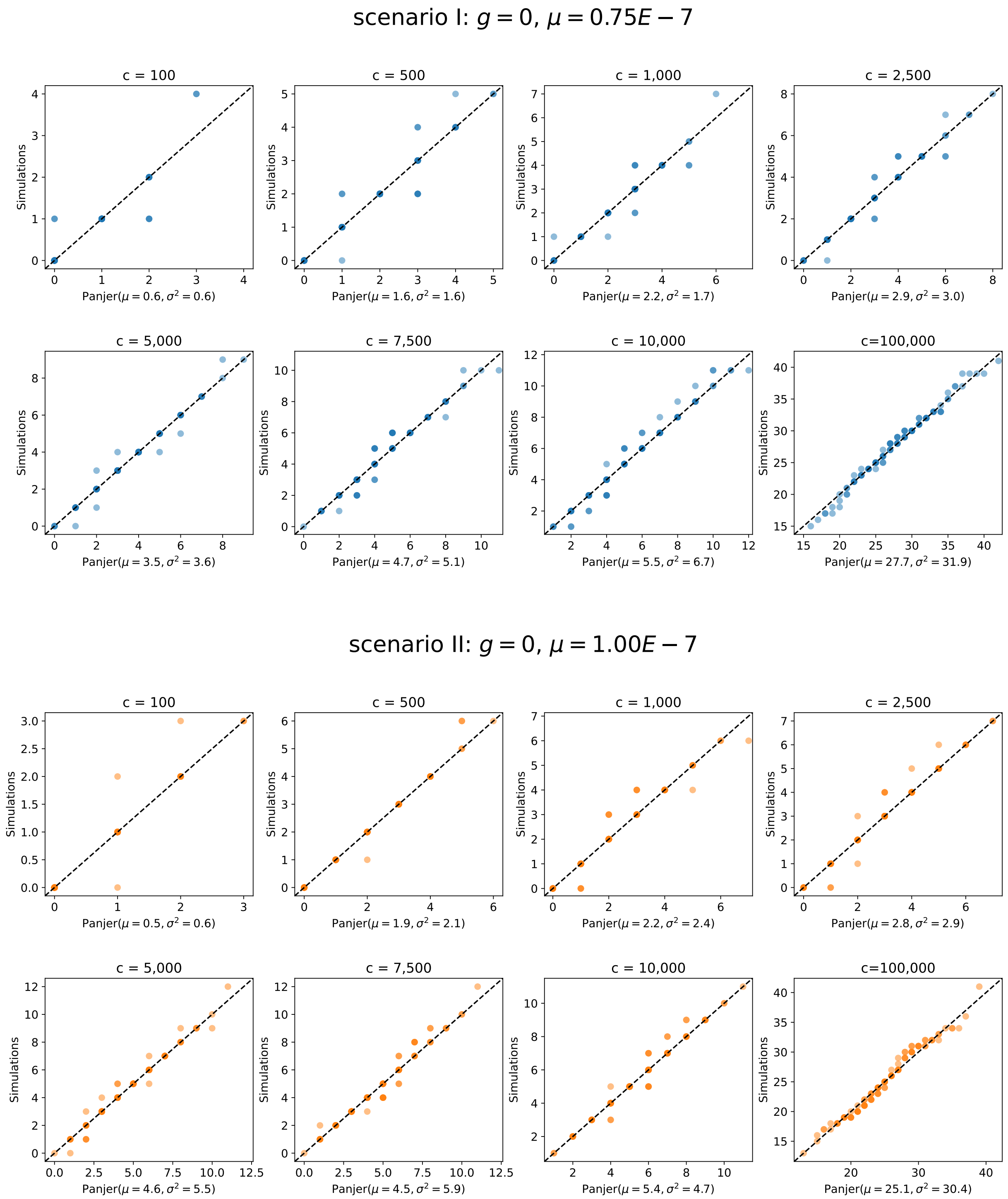

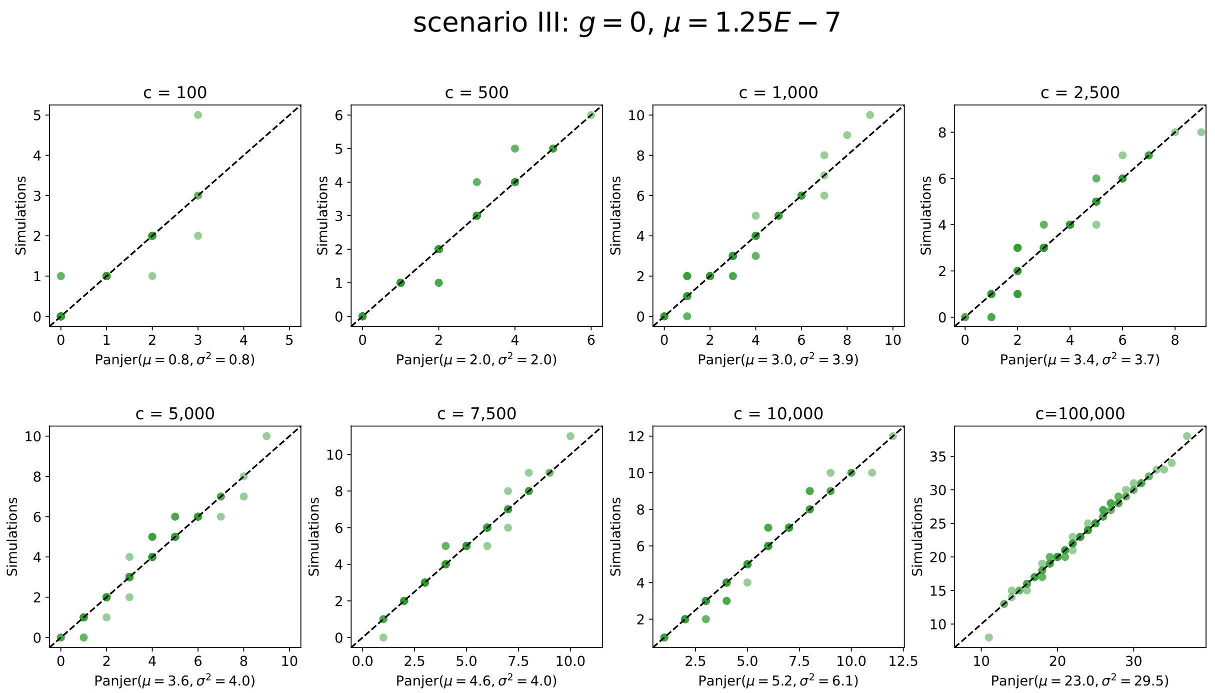
scenario III: $g = 0$, $\mu = 1.25E - 7$
c = 100
Panjer($\mu = 0.8, \sigma^2 = 0.8$)
c = 500
Panjer($\mu = 2.0, \sigma^2 = 2.0$)
c = 1,000
Panjer($\mu = 3.0, \sigma^2 = 3.9$)
c = 2,500
Panjer($\mu = 3.4, \sigma^2 = 3.7$)
c = 5,000
Panjer($\mu = 3.6, \sigma^2 = 4.0$)
c = 7,500
Panjer($\mu = 4.6, \sigma^2 = 4.0$)
c = 10,000
Panjer($\mu = 5.2, \sigma^2 = 6.1$)
c=100,000
Panjer($\mu = 23.0, \sigma^2 = 29.5$)
Simulations

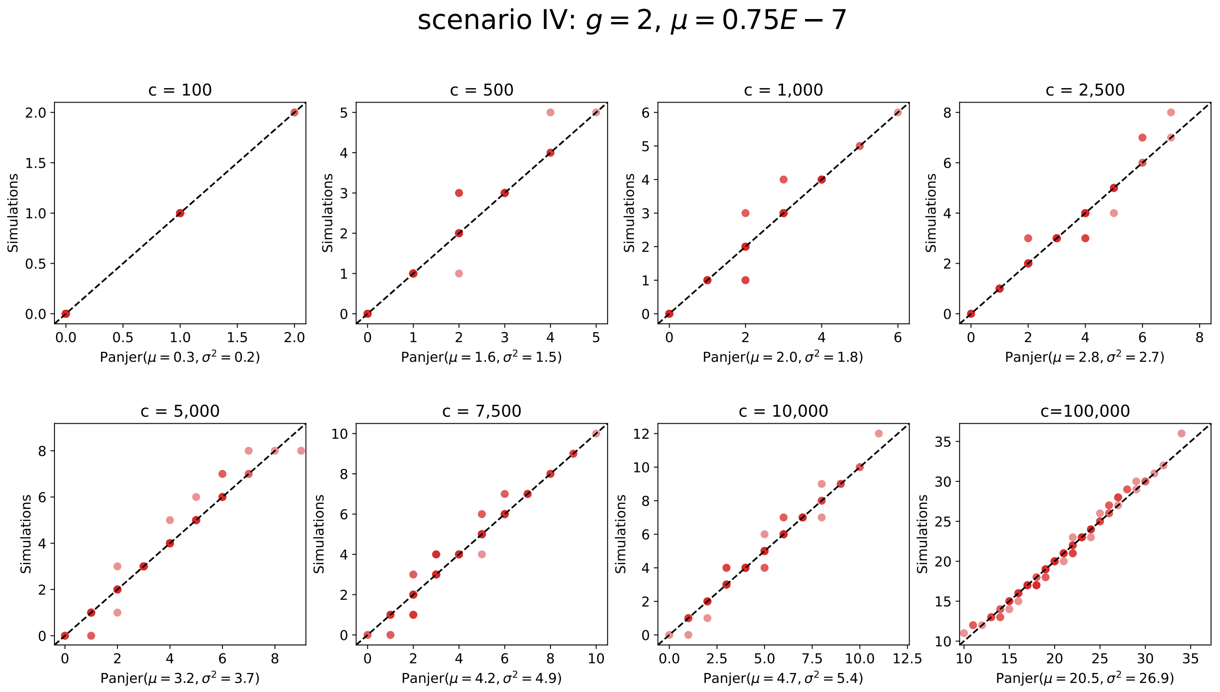
scenario IV: $g = 2$, $\mu = 0.75E - 7$
c = 100
Panjer($\mu = 0.3, \sigma^2 = 0.2$)
c = 500
Panjer($\mu = 1.6, \sigma^2 = 1.5$)
c = 1,000
Panjer($\mu = 2.0, \sigma^2 = 1.8$)
c = 2,500
Panjer($\mu = 2.8, \sigma^2 = 2.7$)
c = 5,000
Panjer($\mu = 3.2, \sigma^2 = 3.7$)
c = 7,500
Panjer($\mu = 4.2, \sigma^2 = 4.9$)
c = 10,000
Panjer($\mu = 4.7, \sigma^2 = 5.4$)
c=100,000
Panjer($\mu = 20.5, \sigma^2 = 26.9$)
Simulations

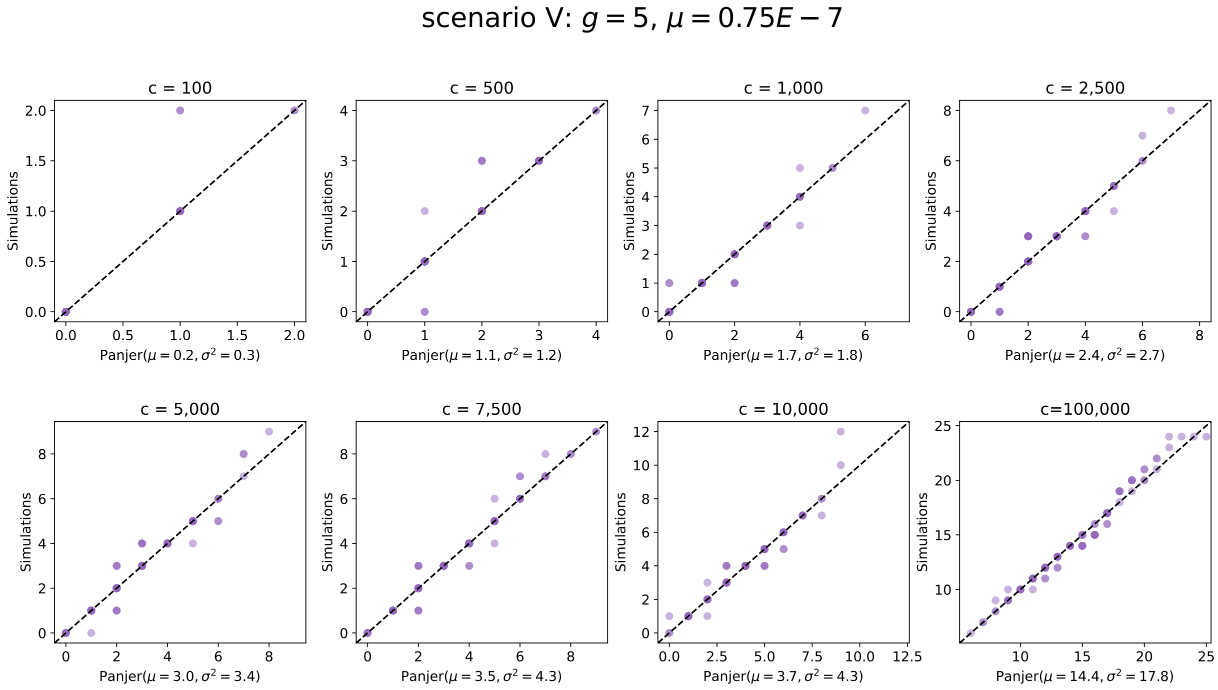
scenario V: $g = 5$, $\mu = 0.75E - 7$
c = 100
c = 500
c = 1,000
c = 2,500
c = 5,000
c = 7,500
c = 10,000
c=100,000
Simulations
Panjer($\mu = 0.2, \sigma^2 = 0.3$)
Panjer($\mu = 1.1, \sigma^2 = 1.2$)
Panjer($\mu = 1.7, \sigma^2 = 1.8$)
Panjer($\mu = 2.4, \sigma^2 = 2.7$)
Panjer($\mu = 3.0, \sigma^2 = 3.4$)
Panjer($\mu = 3.5, \sigma^2 = 4.3$)
Panjer($\mu = 3.7, \sigma^2 = 4.3$)
Panjer($\mu = 14.4, \sigma^2 = 17.8$)

**Figure 2. Analysis of models that extrapolate the simulated data.** For each scenario we fit quadratic, cubic, powerlaw, and exponential models to the observed number of one-dimensional cycles in simulations. Then we fit a linear and powerlaw model to the corresponding residuals as an estimate for the variance of the data. The quality of each model is evaluated through the log-likelihood to observe the validation dataset given a certain model.

#### scenario I: $g = 0, \mu = 0.75E - 7$

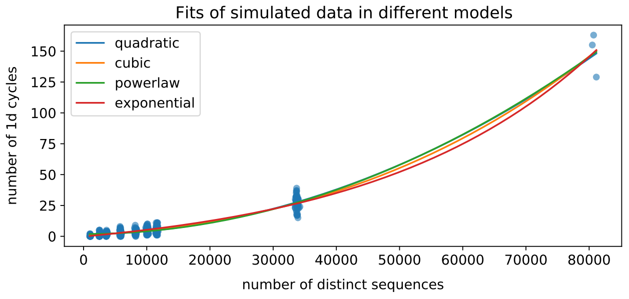


Log-Likelihood of validation data

| | linear | powerlaw |
|---|---|---|
| **quadratic** | -1082 | -665 |
| **cubic** | -1299 | -663 |
| **powerlaw** | -1132 | -669 |
| **exponential** | -1062 | -707 |

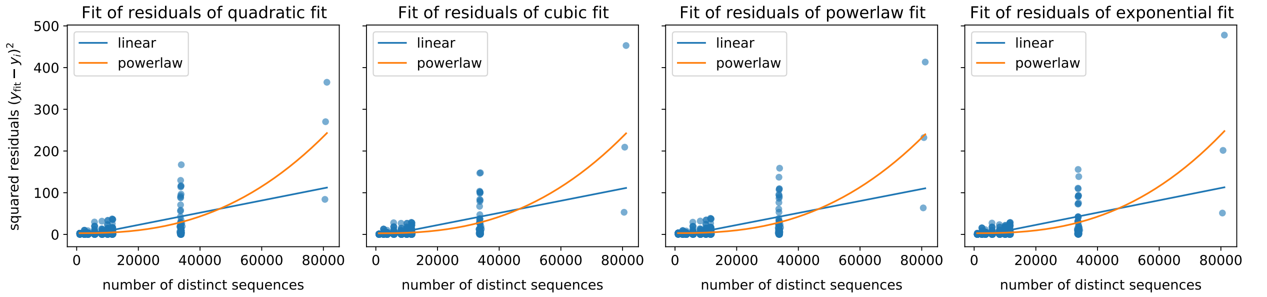


#### scenario II: $g = 0, \mu = 1.00E - 7$

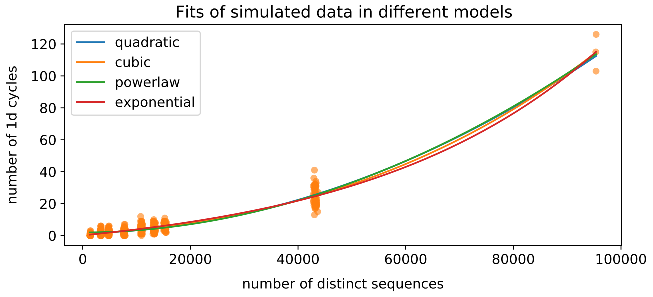


Log-Likelihood of validation data

| | linear | powerlaw |
|---|---|---|
| **quadratic** | -1023 | -685 |
| **cubic** | -831 | -693 |
| **powerlaw** | -1002 | -703 |
| **exponential** | -902 | -653 |

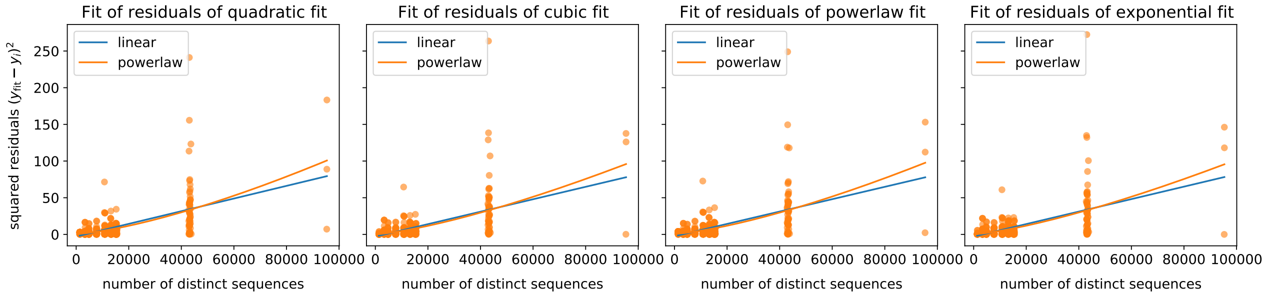

#### scenario III: $g = 0, \mu = 1.25E - 7$

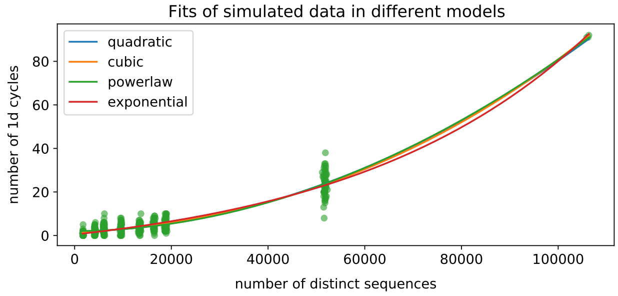


Log-Likelihood of validation data

| | linear | powerlaw |
|---|---|---|
| **quadratic** | -713 | -797 |
| **cubic** | -701 | -714 |
| **powerlaw** | -728 | -730 |
| **exponential** | -689 | -741 |

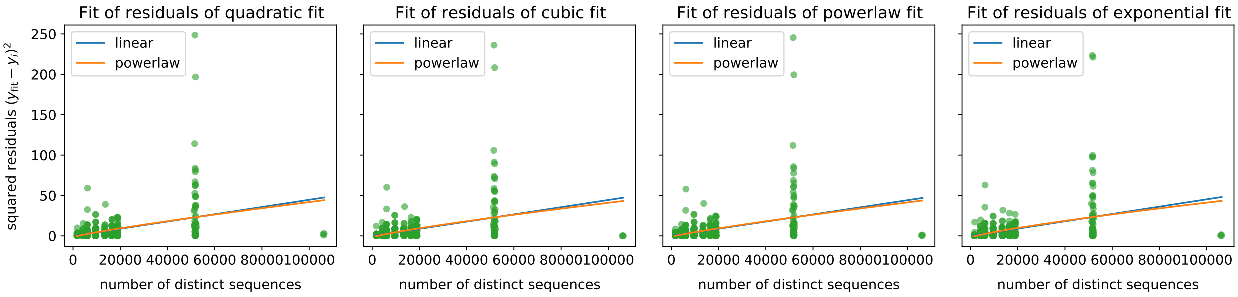


#### scenario IV: $g = 2, \mu = 0.75E - 7$

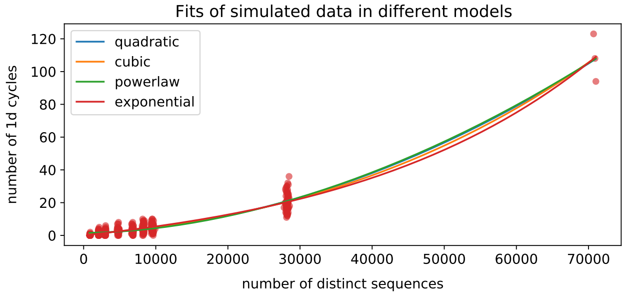


Log-Likelihood of validation data

| | linear | powerlaw |
|---|---|---|
| **quadratic** | -1340 | -638 |
| **cubic** | -981 | -625 |
| **powerlaw** | -1178 | -656 |
| **exponential** | -1018 | -626 |

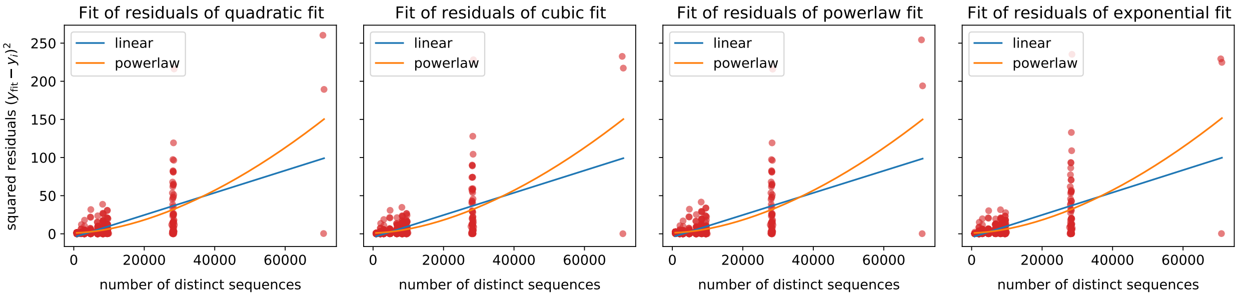

#### scenario V: $g = 5, \mu = 0.75E - 7$

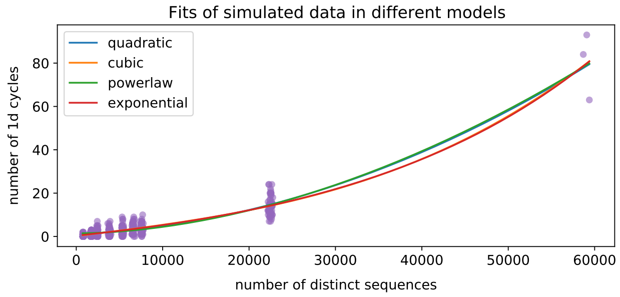


Log-Likelihood of validation data

| | linear | powerlaw |
|---|---|---|
| **quadratic** | -1088 | -602 |
| **cubic** | -875 | -593 |
| **powerlaw** | -1198 | -615 |
| **exponential** | -909 | -593 |

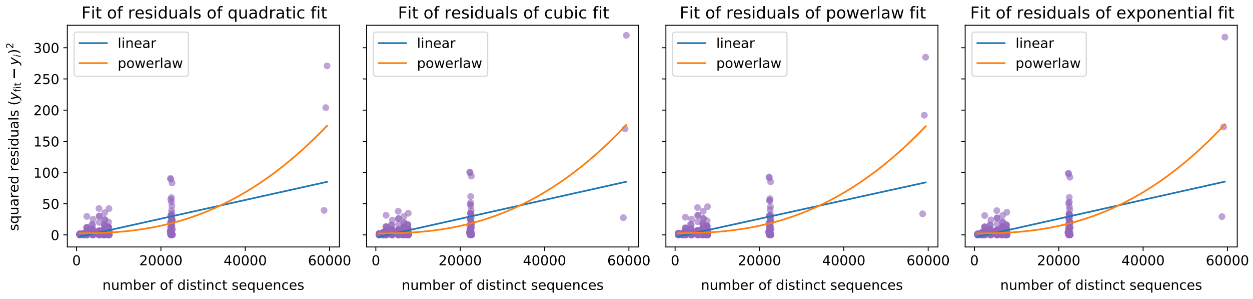

**Figure 3. Comparison of raw counts in SNV cycles and raw tRI.** **(a)** Tables showing the sum of raw counts in SNV cycles and the sum of raw tRI signals, as well as the Pearson correlation coefficients between profiles of raw counts vs. raw tRI signals, for the SARS-CoV-2 (whole genome and spike gene), H5N1 (HA and PB2 gene) and HIV-1 (Env gene) alignments. **(b-c)** Pearson and Spearman correlation coefficients across collection dates for the SARS- CoV-2 whole genome alignment (b) and spike gene alignment (c). **(d-e)** Pearson and Spearman correlation coefficients across collection dates for the H5N1 HA gene alignment (d) and PB2 gene alignment (e). Plots in (b-e) begin when correlations were first defined, and days are counted from the reference sequence date.

**a**

| | | Sum of raw counts in SNV cycles | Sum of raw tRI signals in SNV cycles | Raw counts in SNV cycles vs. raw tRI signals | |
|---|---|---|---|---|---|
| | | | | Pearson r | Spearman ρ |
| SARS-CoV-2 whole genome | day 100 | 41 | 26 | 0.94 | 0.81 |
| | day 500 | 1,453 | 878 | 0.99 | 0.99 |
| | day 1000 | 23,691 | 16,583 | 1.00 | 0.99 |
| | day 1493 | 24,317 | 16,964 | 1.00 | 0.99 |
| SARS-CoV-2 spike gene | day 100 | 37 | 24 | 0.98 | 0.70 |
| | day 500 | 21,743 | 18,131 | 1.00 | 0.99 |
| | day 1000 | 133,819 | 116,003 | 1.00 | 0.99 |
| | day 1492 | 169,638 | 146,060 | 1.00 | 1.00 |
| H5N1 HA gene | day 100 | 400 | 274 | 0.97 | 0.96 |
| | day 500 | 4,553 | 3,022 | 0.98 | 0.98 |
| | day 1000 | 6,504 | 4,322 | 0.98 | 0.99 |
| | day 1410 | 7,827 | 5,172 | 0.98 | 0.99 |
| H5N1 PB2 gene | day 100 | 218 | 131 | 0.97 | 0.93 |
| | day 500 | 3,656 | 2,352 | 0.98 | 0.98 |
| | day 1000 | 5,143 | 3,394 | 0.98 | 0.98 |
| | day 1409 | 5,991 | 3,933 | 0.98 | 0.98 |
| HIV-1 Env gene | overall | 3,931 | 2,523 | 0.97 | 0.98 |

**b**

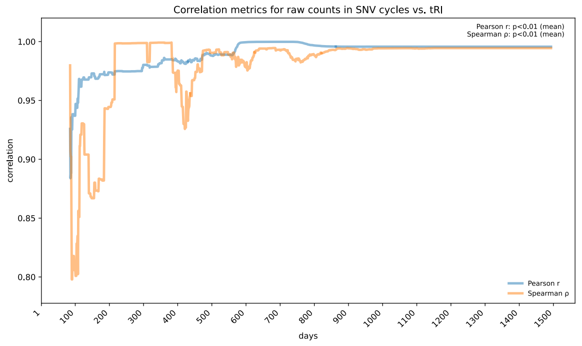


**c**

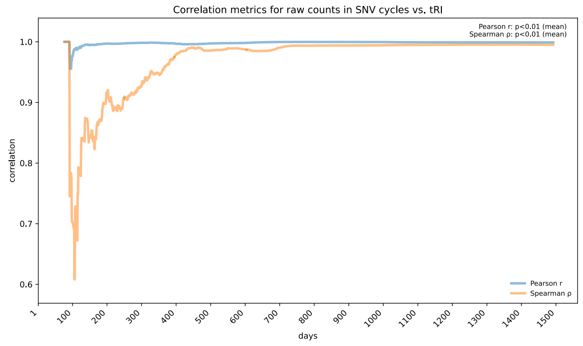


**d**

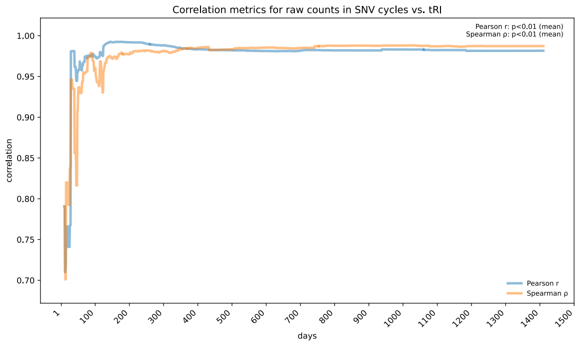


**e**

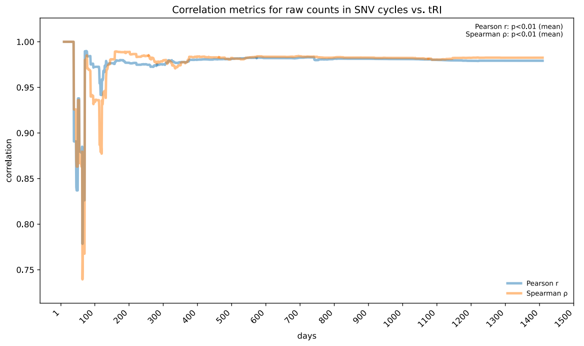

**Figure 4. The effect of missing edges in triangles on raw counts in SNV cycles.** **(a)** Tables showing the sum of (mutation) raw counts in SNV cycles and the sum of signals from missing edges in triangles, as well as the Pearson correlation coefficients between profiles of raw counts vs. combined signals (raw counts plus signals obtained from missing edges in triangles), for the SARS-CoV-2 (whole genome and spike gene), H5N1 (HA and PB2 gene) and HIV-1 (Env gene) alignments. **(b-c)** Pearson and Spearman correlation coefficients across collection dates for the SARS-CoV-2 whole genome alignment (b) and spike gene alignment (c). **(d-e)** Pearson and Spearman correlation coefficients across collection dates for the H5N1 HA gene alignment (d) and PB2 gene alignment (e). Plots in (b-e) begin when correlations were first defined, and days are counted from the reference sequence date.

a

| | | Sum of raw counts in SNV cycles | Sum of signals from missing edges in triangles | Raw counts in SNV cycles vs. combined signal: Pearson r | Spearman ρ |
|---|---|---|---|---|---|
| SARS-CoV-2 whole genome | day 100 | 41 | 0 | 1.00 | 1.00 |
| | day 500 | 1,453 | 12 | 1.00 | 0.99 |
| | day 1000 | 23,691 | 2,942 | 1.00 | 0.95 |
| | day 1493 | 24,317 | 2,946 | 1.00 | 0.95 |
| SARS-CoV-2 spike gene | day 100 | 37 | 9 | 0.97 | 0.97 |
| | day 500 | 21,743 | 1,023 | 1.00 | 0.98 |
| | day 1000 | 133,819 | 5,174 | 1.00 | 0.99 |
| | day 1492 | 169,638 | 5,894 | 1.00 | 0.99 |
| H5N1 HA gene | day 100 | 400 | 2 | 1.00 | 1.00 |
| | day 500 | 4,553 | 78 | 0.99 | 0.99 |
| | day 1000 | 6,504 | 401 | 0.95 | 0.98 |
| | day 1410 | 7,827 | 494 | 0.95 | 0.99 |
| H5N1 PB2 gene | day 100 | 218 | 6 | 0.95 | 0.94 |
| | day 500 | 3,656 | 15 | 1.00 | 1.00 |
| | day 1000 | 5,143 | 622 | 0.92 | 0.99 |
| | day 1409 | 5,991 | 627 | 0.91 | 0.99 |
| HIV-1 Env gene | overall | 3,931 | 167 | 0.96 | 0.98 |

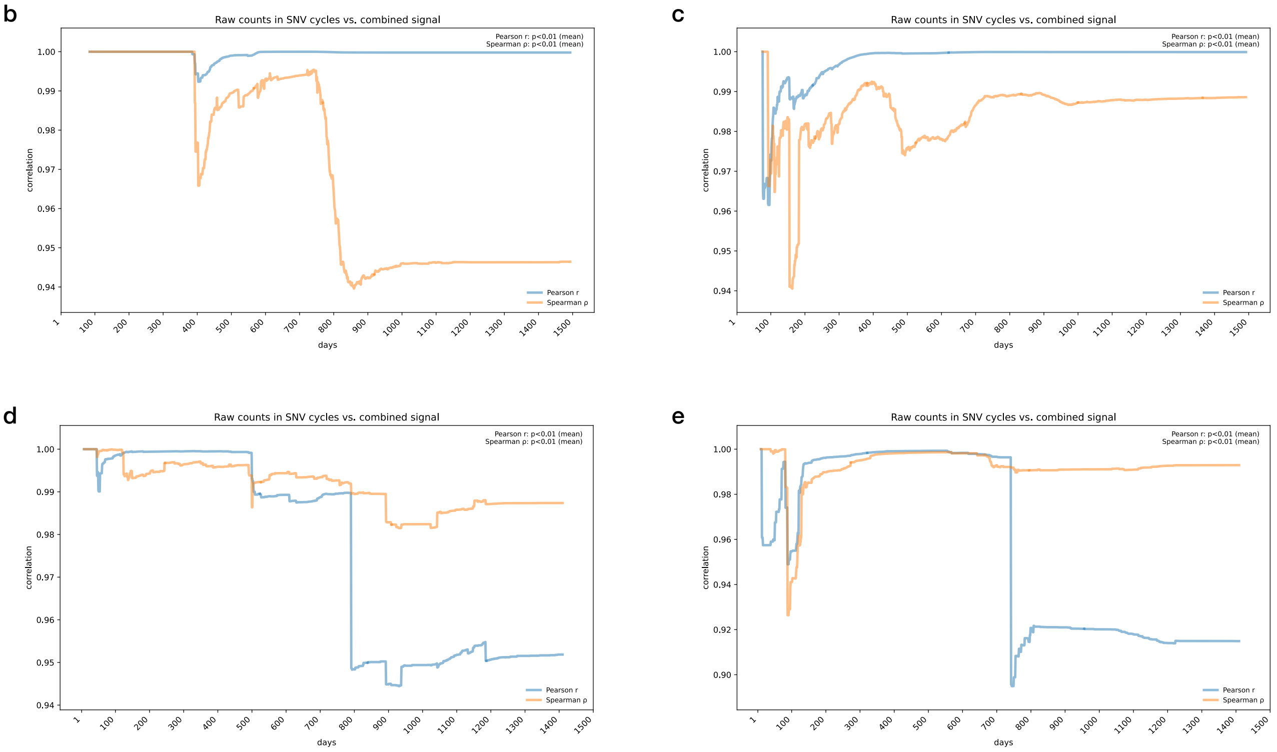

#### References


1. Elbe, S. & Buckland-Merrett, G. Data, disease and diplomacy: GISAID's innovative contribution to global health. *Global Challenges* **1,** 33–46 (2017). doi:10.1002/gch2.1018.
2. Katoh, K. MAFFT: a novel method for rapid multiple sequence alignment based on fast Fourier transform. *Nucleic Acids Research* **30,** 3059–3066 (2002). doi:10.1093/nar/gkf436.
3. Hadfield, J., Megill, C., Bell, S. M., *et al.* Nextstrain: Real-Time Tracking of Pathogen Evolution. *Bioinformatics* **34,** 4121–4123 (2018). doi:10.1093/bioinformatics/bty407.
4. Leinonen, R., Akhtar, R., Birney, E., *et al.* The European Nucleotide Archive. *Nucleic Acids Research* **39,** D28–D31 (2010). doi:10.1093/nar/gkq967.
5. Jariani, A., Warth, C., Deforche, K., *et al.* SANTA-SIM: Simulating Viral Sequence Evolution Dynamics under Selection and Recombination. *Virus Evolution* **5** (2019). doi:10.1093/ve/vez003.
6. Keegan, L. & Kempf, D. *Hammingdist: A Fast Tool to Calculate Hamming Distances* version 1.4.0. 2025. https://github.com/ssciwr/hammingdist visited on 2025-11-01.
7. Bauer, U. Ripser: efficient computation of Vietoris-Rips persistence barcodes. *Journal of Applied and Computational Topology* (2021). doi:10.1007/s41468-021-00071-5.
8. Bauer, U. *Ripser* Github. https://github.com/Ripser/ripser/tree/tight-representative-cycles visited on 2024-01-15.
9. Panjer, H. H. Recursive Evaluation of a Family of Compound Distributions. *ASTIN Bulletin* **12,** 22–26 (1981). doi:10.1017/S0515036100006796.
10. Sundt, B. & Jewell, W. S. Further Results on Recursive Evaluation of Compound Distributions. *ASTIN Bulletin: The Journal of the IAA* **12,** 27–39 (1981). doi:10.1017/S0515036100006802.
11. Sanjuán, R., Moya, A. & Elena, S. F. The distribution of fitness effects caused by single-nucleotide substitutions in an RNA virus. *Proceedings of the National Academy of Sciences* **101,** 8396–8401 (2004). doi:10.1073/pnas.0400146101.
12. Wong, T., Ly-Trong, N., Ren, H., *et al.* IQ-TREE 3: Phylogenomic Inference Software using Complex Evolutionary Models. *EcoEvoRxiv* (2025). doi:10.32942/x2p62n.
13. Sagulenko, P., Puller, V. & Neher, R. A. TreeTime: Maximum-Likelihood Phylodynamic Analysis. *Virus Evolution* **4** (2018). doi:10.1093/ve/vex042.
14. Starr, T. N., Greaney, A. J., Hannon, W. W., *et al.* Shifting mutational constraints in the SARS-CoV-2 receptor-binding domain during viral evolution. *Science* **377,** 420–424 (2022). doi:10.1126/science.abo7896.
15. Starr, T. N., Greaney, A. J., Stewart, C. M., *et al. Deep mutational scanning of the SARS-CoV-2 RBD in variant backgrounds* Github. https://github.com/jbloomlab/SARS-CoV-2-RBD_DMS_variants/blob/main/results/final_variant_scores/final_variant_scores.csv visited on 2024-08-23.
16. Dadonaite, B., Crawford, K. H., Radford, C. E., *et al.* A pseudovirus system enables deep mutational scanning of the full SARS-CoV-2 spike. *Cell* **186,** 1263–1278.e20 (2023). doi:10.1016/j.cell.2023.02.001.
17. Dadonaite, B., Crawford, K. H., Radford, C. E., *et al. Deep mutational scanning of SARS-CoV-2 Omicron BA.1 spike using a barcoded lentiviral platform* Github. https://github.com/dms-vep/SARS-CoV-2_Omicron_BA.1_spike_DMS_mAbs/blob/main/results/muteffects_functional/muteffects_observed.csv visited on 2024-08-23.
18. Dadonaite, B., Brown, J., McMahon, T. E., *et al.* Spike deep mutational scanning helps predict success of SARS-CoV-2 clades. *Nature* **631,** 617–626 (2024). doi:10.1038/s41586-024-07636-1.

19. Dadonaite, B., Brown, J., McMahon, T. E., *et al. Deep mutational scanning of SARS-CoV-2 BA.2 spike* Github. https://github.com/dms-vep/SARS-CoV-2_Omicron_BA.2_spike_ACE2_binding/blob/main/results/summaries/summary.csv visited on 2024-08-23.

20. Bloom, J. D. & Neher, R. A. Fitness effects of mutations to SARS-CoV-2 proteins. *Virus Evolution* **9** (2023). doi:10.1093/ve/vead055.

21. Bloom, J. D. & Neher, R. A. *Fitness effects of SARS-CoV-2 amino-acid mutations estimated from observed versus expected mutation counts* Github. https://github.com/jbloomlab/SARS2-mut-fitness/blob/main/results_gisaid_2024-04-24/aa_fitness/aamut_fitness_by_clade.csv visited on 2025-06-15.

22. Haddox, H. K., Angehrn, G., Sesta, L., *et al.* The mutation rate of SARS-CoV-2 is highly variable between sites and is influenced by sequence context, genomic region, and RNA structure. *Nucleic Acids Research* **53** (2025). doi:10.1093/nar/gkaf503.

23. Haddox, H. K., Angehrn, G., Sesta, L., *et al. Fitness effects of nucleotide and amino acid mutations updated with novel estimates of neutral rates* Github. https://github.com/neherlab/SARS2-mut-fitness-v2/blob/main/results/aamut_fitness/21K_aamut_fitness.csv visited on 2025-06-15.

24. Haddox, H. K., Angehrn, G., Sesta, L., *et al. Fitness effects of nucleotide and amino acid mutations updated with novel estimates of neutral rates* Github. https://github.com/neherlab/SARS2-mut-fitness-v2/blob/main/results/aamut_fitness/BA.2_aamut_fitness.csv visited on 2025-06-15.

25. Haddox, H. K., Angehrn, G., Sesta, L., *et al. Fitness effects of nucleotide and amino acid mutations updated with novel estimates of neutral rates* Github. https://github.com/neherlab/SARS2-mut-fitness-v2/blob/main/results/aamut_fitness/XBB_aamut_fitness.csv visited on 2025-06-15.

26. Dadonaite, B., Ahn, J. J., Ort, J. T., *et al.* Deep mutational scanning of H5 hemagglutinin to inform influenza virus surveillance. *PLOS Biology* **22,** e3002916 (2024). doi:10.1371/journal.pbio.3002916.

27. Dadonaite, B., Ahn, J. J., Ort, J. T., *et al. Deep mutational scanning of H5N1 influenza haemagglutinin using a barcoded lentiviral platform* Github. https://github.com/dms-vep/Flu_H5_American-Wigeon_South-Carolina_2021-H5N1_DMS/blob/main/results/summaries/phenotypes.csv visited on 2025-03-01.

28. Soh, Y. S., Moncla, L. H., Eguia, R., *et al.* Comprehensive mapping of adaptation of the avian influenza polymerase protein PB2 to humans. *eLife* **8** (2019). doi:10.7554/elife.45079.

29. Soh, Y. S., Moncla, L. H., Eguia, R., *et al. Deep mutational scanning of avian influenza PB2 to comprehensively map adaptation to the human host* Github. https://github.com/jbloomlab/PB2-DMS/blob/master/results/diffsel/summary_prefs_effects_diffsel.csv visited on 2025-03-01.

30. Bauer, U. & Roll, F. *Gromov Hyperbolicity, Geodesic Defect, and Apparent Pairs in Vietoris-Rips Filtrations* in (Schloss Dagstuhl – Leibniz-Zentrum für Informatik, 2022). doi:10.4230/LIPICS.SOCG.2022.15.

31. Minh, B. Q., Schmidt, H. A., Chernomor, O., *et al.* IQ-TREE 2: New Models and Efficient Methods for Phylogenetic Inference in the Genomic Era. *Molecular Biology and Evolution* **37,** 1530–1534 (2020). doi:10.1093/molbev/msaa015.

32. Piñeiro, C., Abuín, J. M. & Pichel, J. C. Very Fast Tree: speeding up the estimation of phylogenies for large alignments through parallelization and vectorization strategies. *Bioinformatics* **36** (ed Ponty, Y.) 4658–4659 (2020). doi:10.1093/bioinformatics/btaa582.

33. Piñeiro, C. & Pichel, J. C. Efficient phylogenetic tree inference for massive taxonomic datasets: harnessing the power of a server to analyze 1 million taxa. *GigaScience* **13** (2024). doi:10.1093/gigascience/giae055.

34. De Maio, N., Kalaghatgi, P., Turakhia, Y., *et al.* Maximum likelihood pandemic-scale phylogenetics. *Nature Genetics* **55,** 746–752 (2023). doi:10.1038/s41588-023-01368-0.

35. Ly-Trong, N., Bielow, C., De Maio, N. & Minh, B. Q. CMAPLE: Efficient Phylogenetic Inference in the Pandemic Era. *Molecular Biology and Evolution* **41** (ed Rzhetsky, A.) (2024). doi:10.1093/molbev/msae134.

36. Turakhia, Y., Thornlow, B., Hinrichs, A. S., *et al.* Ultrafast Sample placement on Existing tRees (UShER) enables real-time phylogenetics for the SARS-CoV-2 pandemic. *Nature Genetics* **53,** 809–816 (2021). doi:10.1038/s41588-021-00862-7.
